# Nocturia as a Risk Factor for Developing Frailty in Older Adults: Results of the Berlin Aging Study II

**DOI:** 10.1101/2024.09.20.24313292

**Authors:** Maximilian König, Carolin Malsch, Joany Mariño, Valentin Max Vetter, Yulia Komleva, Ilja Demuth, Elisabeth Steinhagen-Thiessen

## Abstract

**Background & Aim:** The current study examined cross-sectional and longitudinal associations between nocturia and frailty in a cohort of men and women aged 60 years and older, as evidence on this topic was lacking.

**Methods:** Baseline and follow-up data from the Berlin Aging Study II (n=1671) assessed on average 7.1 (IQR 6.2-8.7) years apart were analyzed. Self-reported nocturia was dichotomized into </≥ 2 micturitions per night, and frailty was assessed using the Fried Frailty Phenotype. Covariables were identified a priori based on a review of the existing literature.

**Results:** At baseline, 70.2% of the participants were robust, 28.9% were pre-frail, and 0.9% were frail; 254 participants (23.6%) had self-reported nocturia. In longitudinal analyses, the prevalence and incidence of frailty at follow-up significantly increased when nocturia was present at baseline. Over the median follow-up of 7.1 years, there were 41 incident frailty cases (IR 5.15, 95% CI 3.79-7.00 per 1000 person-years). After adjusting for age, sex, morbidity burden, and baseline frailty status, baseline nocturia was associated with a 2.23-fold increased risk (95% CI 1.17-4.18) of frailty at follow-up.

**Conclusion:** Nocturia is linked to a higher risk of developing frailty in older adults, both women and men.

## 1. Introduction

While good sleep is pivotal for maintaining normal physical and mental health, disrupted sleep is among older adults’ most common health complaints [1]. Aging is associated with a reduced ability to initiate and maintain sleep—often wrongfully regarded as a feature of ‘normal aging’ [2]. One of the main causes of sleep disturbance in older adults is nocturia [3]. Nocturia is when a person has to interrupt sleep during the night to urinate [4]. Few younger adults report nocturia (< 5%), whereas approximately half of older adults aged 60 years and older and approximately 80% of those aged 80 years and older are affected [5]. Ultimately, the causes of nocturia are unclear: nocturnal polyuria, cardiovascular disease and related (poly)pharmacotherapy, reduced bladder capacity, sleep disturbances, insomnia, or a combination of these and other factors are often thought to be responsible [6], [7]. Nocturia can have a major impact on overall health and quality of life [3]. For example, it can put older adults at increased risk of falling if they get up to urinate in the dark. More importantly, it disturbs sleep and has a manifold of negative consequences. Consequently, nocturia is associated with daytime dysfunction and sleepiness, fractures, reduced gait speed, reduced functionality, and mobility, among others [2], [4]; moreover, two or more nocturia episodes have been shown to be a marker of poor health in older women [8]. With demographic change, the proportion of old and very old people continues to increase. Unfortunately, healthspan is not keeping pace with the increase in life expectancy, so age-associated diseases, geriatric syndromes, and frailty are becoming increasingly important [9],[10],[11]. Frailty is a clinically identifiable state of diminished physiological reserve and increased vulnerability to a broad range of adverse health outcomes [12]. While the prevalence is increasing, the reasons for the development of frailty are not yet known with sufficient certainty, and strategies to prevent and treat frailty are needed [10].

Healthy, restorative sleep can be considered the foundation of a sustainable healthy lifestyle, given its many essential physiological functions, including cell regeneration, immune function, metabolic function, hormonal function, and cognition [1]. Sleep deprivation affects almost every physiological function in the body and has been associated with reduced life expectancy, accelerated aging, neurodegenerative diseases, dementia, loss of energy and activity, and frailty [13]. Nevertheless, sleep and nocturia are often under-recognized in everyday clinical practice, research, and public health agendas [1]. In particular, their relationship to frailty is not acknowledged. The existing evidence on the association between nocturia and frailty is scarce and contradictory [14]. Some cross-sectional studies have found evidence for an association, but there are often design, methods, and analysis constraints, limiting their value to disentangling the complex relationship [14]. Studies that would allow causal inference, e.g., longitudinal studies, are still missing.

Using longitudinal data from the Berlin Aging Study II, we aimed to examine cross-sectional and longitudinal associations between nocturia, frailty, and mortality. We hypothesized that nocturia is a modifiable risk factor for incident frailty.

## 2. Methods

### 2.1. Study population and design

The Berlin Aging Study II (BASE-II) is a prospective cohort study investigating factors associated with “healthy” and “unhealthy” aging, and has been described in detail previously [15], [16]. Participants were community-dwelling, comparably well-functioning older adults (aged between 60 and 84 years), and a control group of young adults. The baseline medical assessments took place between 2009 and 2014. Follow-up data from 1083 BASE-II participants were obtained as part of the GendAge study [17] between 2018 and 2020, and were complemented by information on vital status received from the civil register. Our analysis used data from the baseline assessments of the older age group in BASE-II and the corresponding follow-up assessments in GendAge.

The study was approved by the ethics committee of the Charité-Universitätsmedizin Berlin (EA2/029/09 and EA2/144/16) and conducted according to the declaration of Helsinki. All participants gave written informed consent for study participation.

### 2.2 Nocturia assessment

Nocturia was assessed using the same two questions, both at baseline and follow-up: “Do you have to get up at night to urinate?” and “How often do you need to urinate per night?” We operationalized nocturia as two or more episodes of nocturia. [18].

### 2.3 Mortality

The vital status was periodically checked at the civil register, most recently in November 2023.

### 2.4 Frailty assessment

Frailty was operationalized according to the phenotype concept proposed by Fried et al. [19]. As previously described, minor modifications to the original methodology were necessary to adjust for small differences in how the criteria were assessed in BASE-II [20]. The five frailty phenotype criteria were assessed as follows:

- “Unintentional weight loss”: Unintentional loss of at least 5% of body weight during the past year.
- “Self-reported exhaustion”: two questions from the “Center for Epidemiological Studies depression” scale [21].
- “Weakness”: low handgrip strength measured by Smedley Dynamometer (Scandidact, Denmark). We used the original cut-off values as suggested by Fried et al. [19].
- “Slow walking speed”: walking speed was assessed in the Timed Up & Go test [22].
- “Low physical activity”: was assessed with the question, “Are you seldom or never physically active?”

According to the number of components present, participants were categorized as frail (3–5), prefrail (1–2), or robust (0)[19].

### 2.5 Covariables

Sleep was assessed using the Pittsburgh Sleep Quality Index, a widely used tool in clinical practice and research for assessing subjective sleep habits and quality [23]. The questionnaire consists of 19 questions evaluating subjective sleep quality, latency to fall asleep, sleep duration, sleep efficiency, sleep disorders, use of sleep medication, and impairment of daytime activity. Regarding subjective sleep quality, we grouped the categories “poor” and “very poor” to create the variable poor subjective sleep quality. Unfortunately, the subjective sleep quality question was unavailable at follow-up.

All other diagnoses were recorded in a structured manner as part of the data collection process, integrating information from various sources. These were used to compute a morbidity index largely based on the categories of the Charlson Comorbidity Index, which is a weighted sum of moderate to severe, mostly chronic physical illnesses, including cardiovascular diseases (e.g., congestive heart failure), cancer (e.g., lymphoma), and metabolic diseases (e.g., diabetes mellitus) [24], [25]. Further self-reported information included in the analysis were falls and hospitalization in the past 12 months prior to the baseline and follow-up assessments, respectively.

### 2.5 Statistical analyses

Participant’s characteristics were compared between strata using a t-test, one-way analysis of variance (ANOVA), Kruskal‒Wallis test, Wilcoxon rank sum test, Fisher’s test, or χ^2^ test, as appropriate.

The main outcomes of interest were frailty and mortality, while the exposure was nocturia. Hence, we calculated Kaplan-Meier curves and mortality rates per 1000 person-years (PY) with 95% percentile bootstrapping confidence intervals, stratified by nocturia and frailty status at baseline. Incidence rates of frailty per 1000 PY with percentile bootstrapping confidence intervals to a level of 95% were calculated stratified for baseline nocturia.

As the duration of follow-up was heterogeneous between participants, we analyzed the relationship between nocturia and incident frailty through (1) a Poisson regression with the duration of observation as an offset in the regression function, and (2) adjusting for the duration of observation in the logistic regression (among other confounders). Incidence rate ratios (IRRs) and odds ratios (ORs) with 95% bootstrapping confidence intervals (95% CIs) are presented after adjusting for key confounders (age, sex, frailty status at baseline, and morbidity index). Frailty transitions were visualized in an alluvial plot. A comprehensive assessment of the predictors of frailty was beyond this study’s aim; thus, we limited our models to including the variables mentioned above.

Participants, who were lost to follow-up and those with missing data for the key exposures and outcomes were not included in the longitudinal analyses with frailty as the outcome.

All analyses were performed using Stata SE 18.0 (StataCorp, College Station, TX) and R (version 4.3.1, R Core Team, 2023). The alluvial plot was created with the ggplot2 [27] and ggalluvial packages [28].

### 2.6 STROBE Statement

The manuscript was prepared in compliance with the STrengthening the Reporting of Observational Studies in Epidemiology (STROBE) statement [29].

## 3. Results

### 3.1 Study population

**Figure 1** illustrates the sampling procedure. The original BASE-II sample included 1671 participants aged 60 years and older (range: 60–84 years). In the follow-up, 1083 participants (64.8%) of the original BASE-II sample were seen at a median of 7.1 years (IQR 6.2-8.7; range: 3.91-10.37 years) after the baseline examination (**Supplementary figure 1**). 126 participants (7.5%) died between baseline and follow-up. 462 participants (27.6%) were lost to follow-up, but were alive according to information from the civil register by March 3, 2023 (end of follow-up assessments). A further 110 participants died after follow-up, giving a total of 236 participants (14.1%) who had died by May 2023. The characteristics of the full baseline sample and the reduced subsample with available follow-up data are presented in **Table 1**.

**Figure 1.**
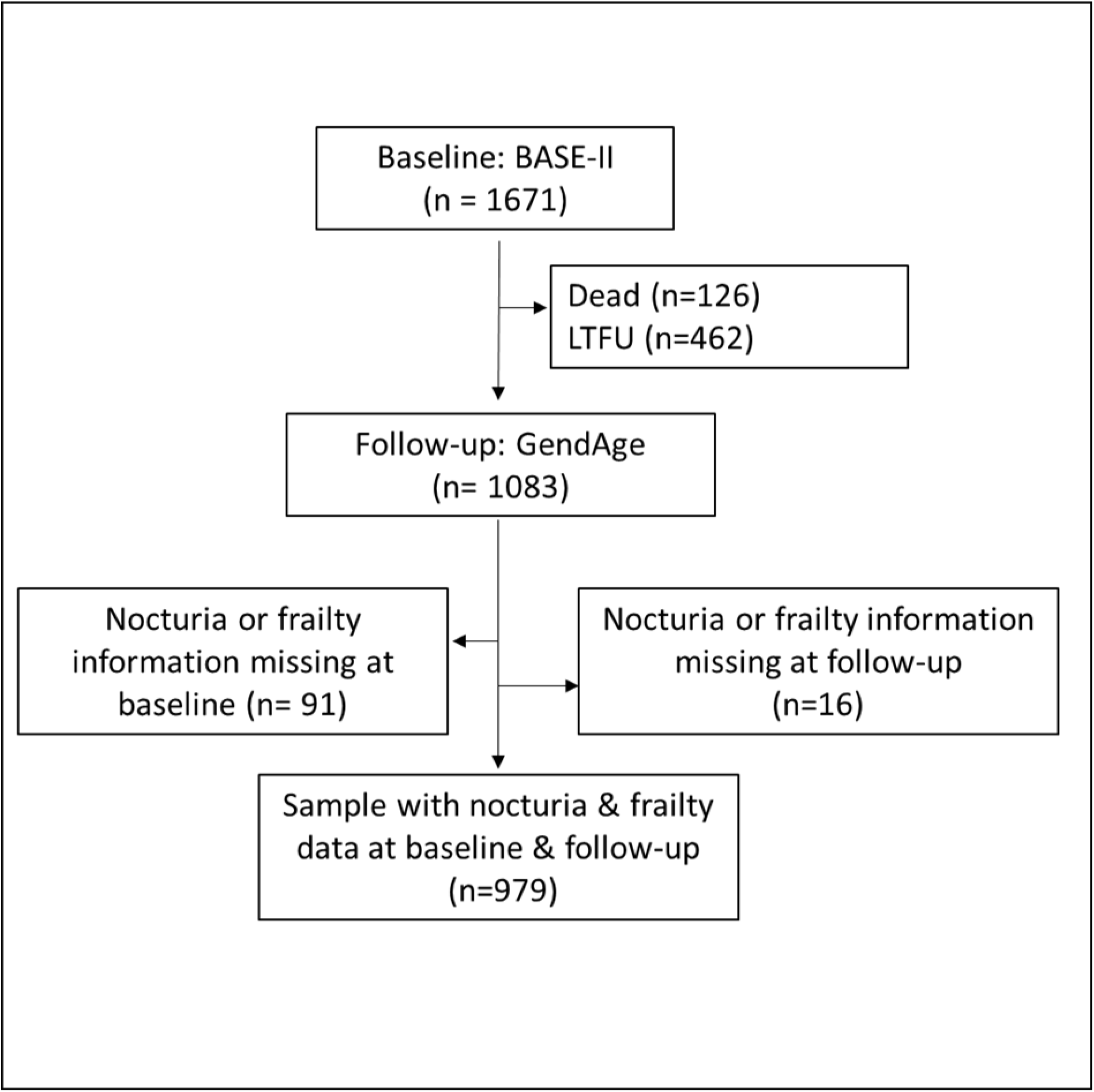
Flowchart showing the derivation of the study population. *GendAge is a subsample of BASE-II participants. BASE-II, Berlin Aging Study II, LTFU,loss to follow-up.

**Table 1.**
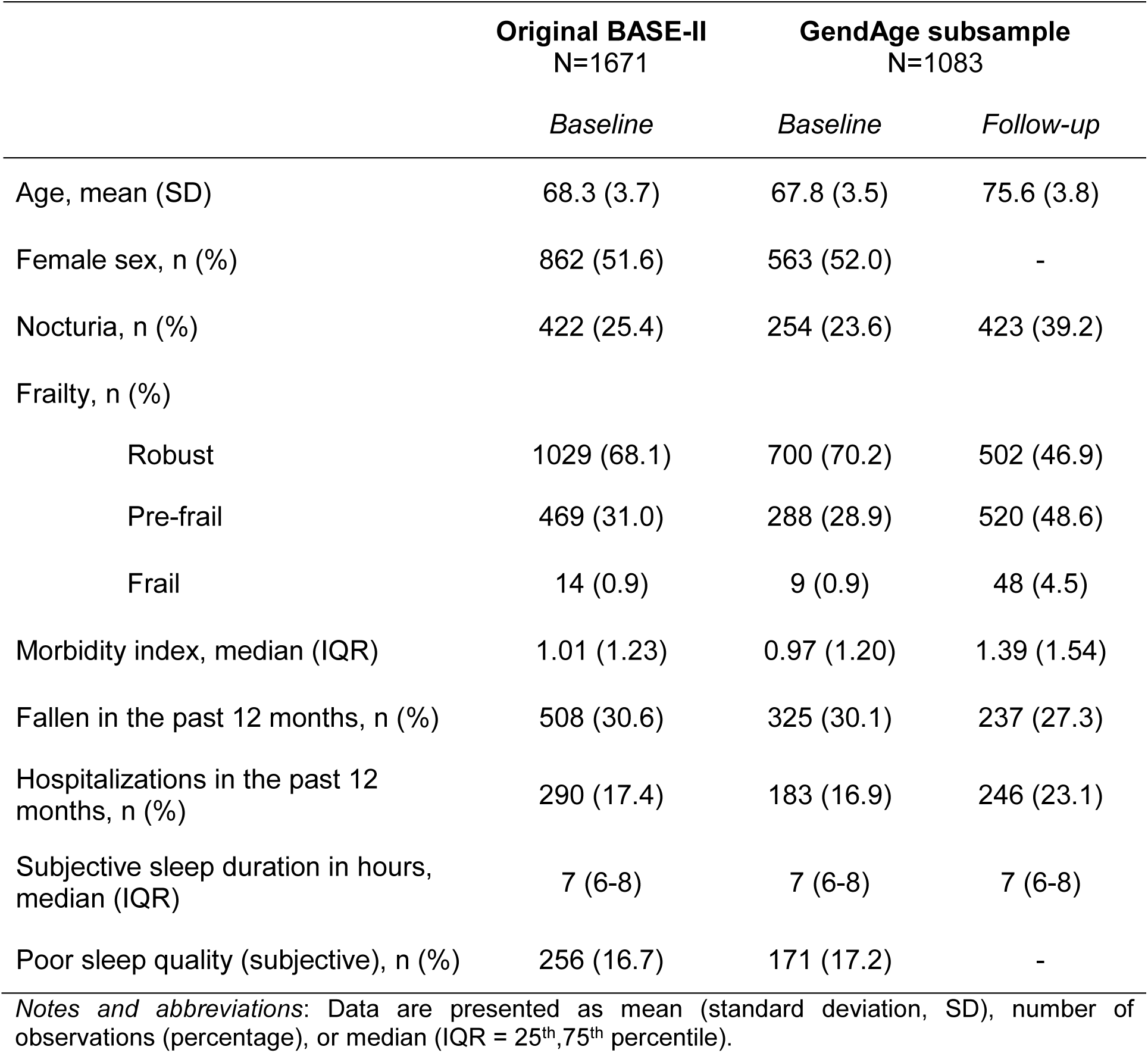
Characteristics of participants in the BASE-II study and the GendAge subsample.

|  | Original BASE-II<br>N=1671 | GendAge subsample<br>N=1083 |  |
| --- | --- | --- | --- |
|  | *Baseline* | *Baseline* | *Follow-up* |
| Age, mean (SD) | 68.3 (3.7) | 67.8 (3.5) | 75.6 (3.8) |
| Female sex, n (%) | 862 (51.6) | 563 (52.0) | - |
| Nocturia, n (%) | 422 (25.4) | 254 (23.6) | 423 (39.2) |
| Frailty, n (%) |  |  |  |
| Robust | 1029 (68.1) | 700 (70.2) | 502 (46.9) |
| Pre-frail | 469 (31.0) | 288 (28.9) | 520 (48.6) |
| Frail | 14 (0.9) | 9 (0.9) | 48 (4.5) |
| Morbidity index, median (IQR) | 1.01 (1.23) | 0.97 (1.20) | 1.39 (1.54) |
| Fallen in the past 12 months, n (%) | 508 (30.6) | 325 (30.1) | 237 (27.3) |
| Hospitalizations in the past 12 months, n (%) | 290 (17.4) | 183 (16.9) | 246 (23.1) |
| Subjective sleep duration in hours, median (IQR) | 7 (6-8) | 7 (6-8) | 7 (6-8) |
| Poor sleep quality (subjective), n (%) | 256 (16.7) | 171 (17.2) | - |

*Notes and abbreviations:* Data are presented as mean (standard deviation, SD), number of observations (percentage), or median (IQR = 25<sup>th</sup>, 75<sup>th</sup> percentile).

Participants not seen at the follow-up (n = 588, 35.2%) were older, more often frail or pre-frail, and more likely to suffer from nocturia at baseline (**Supplementary Table 1**).

### 3.2 Associations between frailty, nocturia, and mortality

The mortality rate per 1000 PY was 12.6 (95%-CI 11.1-14.2). Stratification by baseline nocturia and frailty status revealed significant differences in mortality rates. Mortality was significantly higher in participants with nocturia compared to those without, and in participants with frailty or pre-frailty compared to robust participants (**Table 2a, Supplementary Figures 2a-b**).

**Table 2a.**
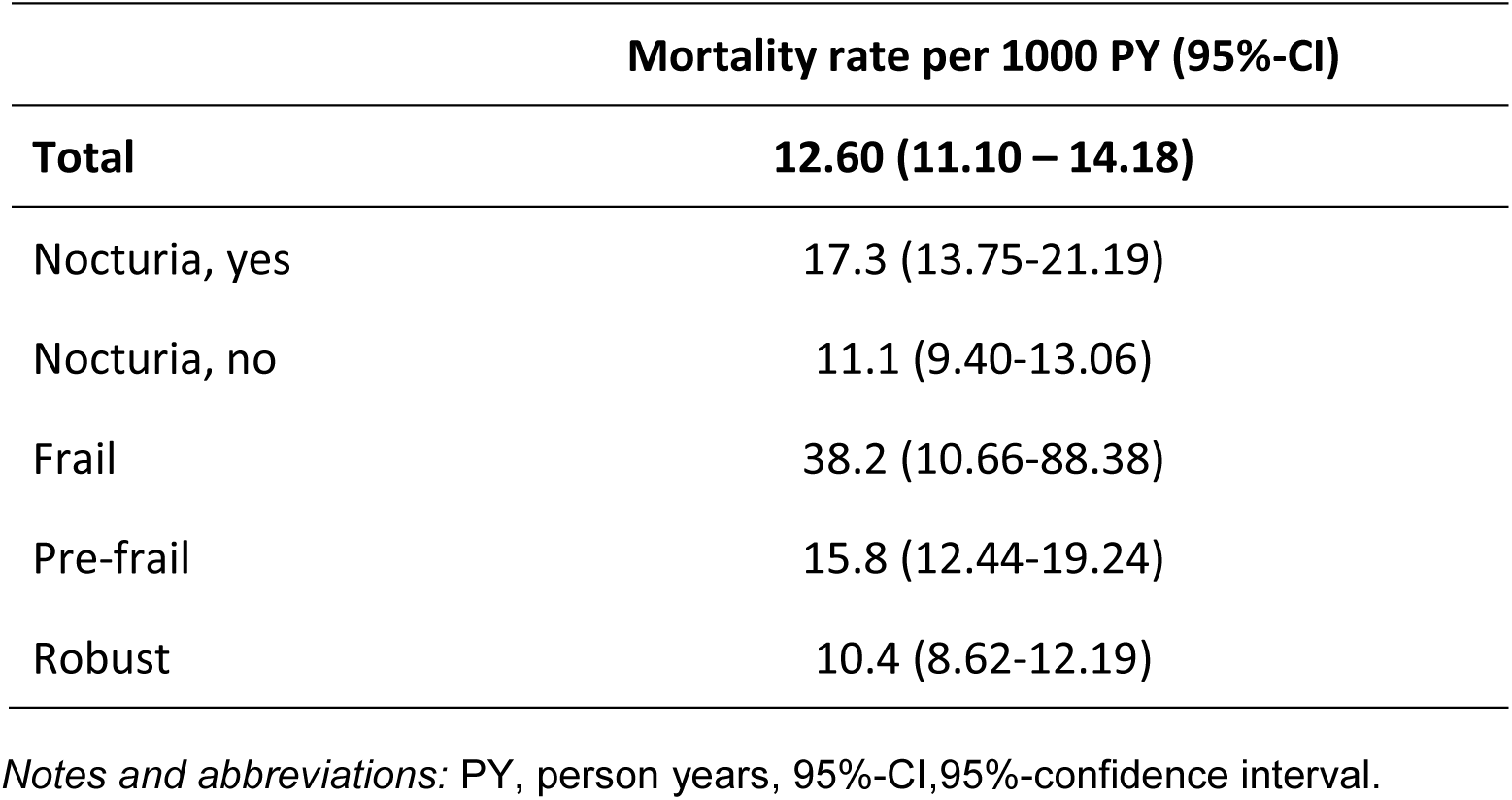
Mortality rates per 1000 person-years (PY), total and stratified by baseline nocturia and frailty status at baseline (n=1671).

|  | Mortality rate per 1000 PY (95%-CI) |
| --- | --- |
| **Total** | **12.60 (11.10 – 14.18)** |
| Nocturia, yes | 17.3 (13.75-21.19) |
| Nocturia, no | 11.1 (9.40-13.06) |
| Frail | 38.2 (10.66-88.38) |
| Pre-frail | 15.8 (12.44-19.24) |
| Robust | 10.4 (8.62-12.19) |

*Notes and abbreviations:* PY, person years, 95%-CI, 95%-confidence interval.

In multivariable analysis, frailty at baseline was significantly associated with death (**Table 2b**). Further significant associations were found for age and sex, but not for pre-frailty, nocturia, and overall morbidity burden.

**Table 2b.** Multivariable-adjusted^#^ Rate Ratios and Odds Ratios of death in BASE-II (n=1671).

|  | Rate Ratio (95%-CI) | Odds Ratio (95%-CI) |
| --- | --- | --- |
| Pre-frail* | 1.42 (0.74-2.66) | 1.44 (0.73-2.77) |
| Frail* | **8.80 (1.40-30.30)** | **8.89 (1.20-43.90)** |
| Nocturia** | 0.69 (0.30-1.42) | 0.68 (0.29-1.44) |
| Baseline age | **1.13 (1.04-1.22)** | **1.14 (1.05-1.24)** |
| Female sex | **0.41 (0.21-0.76)** | **0.44 (0.22-0.86)** |
| Morbidity index | 0.89 (0.67-1.14) | 0.87 (0.64-1.13) |
| Follow-up duration | - | 0.94 (0.75-1.18) |

*Notes and abbreviations:* \* reference: robust, \*\*reference: no nocturia, #adjusted for baseline age, sex, morbidity index and follow-up duration.

### 3.3 Associations between nocturia and frailty

Nocturia prevalence increased from 25.4% at baseline to 39.2% at follow-up (**Table 1**). Prevalence of pre-frailty and frailty increased from 31.0% and 0.9% to 48.6% and 4.5%, respectively. At follow-up, 350 (35.6%) participants had progressed in their frailty status, with 41 (4.2%) new cases of frailty. Figure 2 illustrates the transitions in frailty status from baseline to follow-up stratified by nocturia status. Nocturia was associated with higher probabilities of transitioning from robust to pre-frailty (47.7% vs. 44.6%) and frailty (7.1% vs. 1.3%), and a lower probability of remaining robust (47.7% vs. 54.1%) by follow-up. Likewise, nocturia reduced the likelihood of returning to robust (27.5% vs. 34.4%), while increasing the risk of transitioning to frailty (13.0% vs. 6.6%).

**Figure 2.**
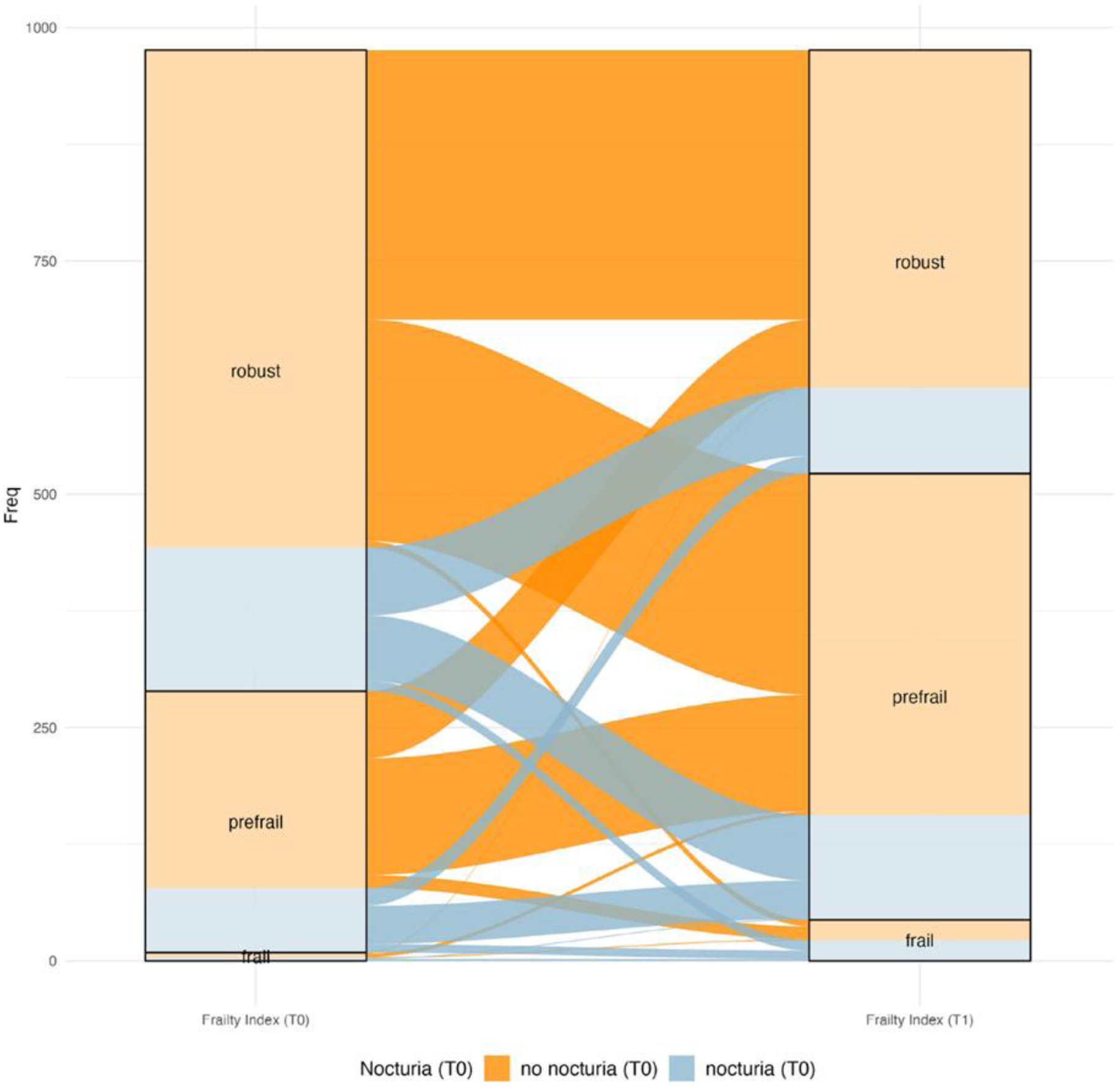
Alluvial plot showing frailty transitions (robust ↔ pre-frail ↔ frail) stratified by nocturia status at baseline (n = 979). At baseline, 689 (70.4%) were robust, 281 (28.7%) pre-frail, and 9 (0.9%) frail. By follow-up, 456 (46.6%) were robust, 479 (48.9%) were pre-frail, and 44 (4.5%) were frail. Of those initially robust, 308 (44.7%) transitioned to pre-frailty, 18 (2.6%) to frailty, while 363 (52.7%) stayed robust. Nocturia was associated with higher probabilities of transitioning to pre-frailty (45.2% vs. 44.6%) and frailty (7.1% vs. 1.3%), and a lower probability of remaining robust (47.7% vs. 54.1%) by follow-up. Among the 281 initially pre-frail participants, 92 (32.7%) returned to robust, 166 (59.1%) remained pre-frail, and 23 (8.2%) progressed to frailty. Nocturia reduced the probability of becoming robust (27.5% vs. 34.4%), while increasing the probability of transitioning to frailty (13.0% vs. 6.6%). Of the 9 participants who were initially frail, 1 (11.1%) returned to robust, 5 (55.6%) became pre-frail, and 3 (33.3%) remained frail at follow-up. Of the 44 frail participants at follow-up, 18 (40.9%) had been robust and 23 (52.3%) pre-frail at baseline. Notably, among 41 new frailty cases, nocturia was present in 11 (61.1%) of those initially robust and 9 (39.1%) of those initially pre-frail.

A cross-sectional analysis revealed no significant association between nocturia and frailty at baseline (p = 0.633), whereas a significant association was identified at follow-up, where frailty and pre-frailty were significantly more prevalent in participants with nocturia than in those without (5.8% vs. 3.7%, and 53.3% vs. 45.7%, respectively; p = 0.005). Longitudinally, participants with nocturia at baseline showed a significantly higher frailty incidence rate (7.4, 95%-CI 4.5-10.8 per 1000 PY) than participants without (2.4, 95%-CI 1.4-3.4, per 1000 PY). According to multivariable Poisson regression adjusted for baseline age, sex, morbidity index, and baseline frailty status, nocturia at baseline was associated with a 2.23-fold increased frailty rate per 1000 PY at follow-up (95%-CI 1.17-4.18, **Table 3**). This result was consistent with multivariable logistic regression, which showed 2.45-fold (95%-CI 1.23-4.83) increased odds of frailty at follow-up when nocturia was present at baseline. Similar results were observed for frailty incidence, with a 2.07-fold increased frailty incidence rate per 1000 PY (95%-CI 1.05-4.00) and 2.23-fold increased odds of incident frailty (95%-CI 1.10-4.43) for participants with nocturia at baseline compared to those without nocturia.

**Table 3.** Associations between nocturia at baseline and follow-up frailty (n = 915).

|  | Rate ratio (95%-CI) | Odds Ratio (95%-CI) |
| --- | --- | --- |
| **Frailty at follow-up** |  |  |
| Nocturia | 2.23 (1.17-4.18) | 2.45 (1.23-4.83) |
| Age at baseline | 1.11 (1.02-1.20) | 1.12 (1.02-1.22) |
| Female Sex | 1.21 (0.65-2.31) | 1.14 (0.57-2.32) |
| Morbidity index at baseline | 1.42 (1.16-1.72) | 1.53 (1.22-1.90) |
| Frailty at baseline | 3.26 (1.86-5.65) | 3.89 (2.12-7.26) |
| Follow-up duration | - | 1.33 (1.04-1.70) |
| **Incident Frailty at follow-up** |  |  |
| Nocturia | 2.07 (1.05-4.00) | 2.23 (1.10-4.43) |
| Age at baseline | 1.13 (1.03-1.23) | 1.14 (1.04-1.26) |
| Female Sex | 1.32 (0.69-2.58) | 1.27 (0.63-2.61) |
| Morbidity index at baseline | 1.53 (1.24-1.86) | 1.62 (1.29-2.03) |
| Follow-up duration | - | 1.29 (1.01-1.66) |

*Notes and abbreviations:* 95%-CI,95% confidence interval.

To complete the emerging clinical picture, the clinical characteristics of participants at both time points and stratified by nocturia and frailty status are presented in **Supplementary Tables 2 and 3**. Participants with nocturia were consistently older, had higher hospitalization rates, and exhibited more overall morbidity than those without nocturia at both time points. No significant sex difference was observed at baseline; however, at follow-up, nocturia was more prevalent among male participants. Self-reported sleep duration was shorter in people with nocturia, and more people with nocturia reported poor sleep quality at baseline.

## 4. Discussion

Our cross-sectional and longitudinal analyses of the Berlin Aging Study II data suggest that nocturia is associated with frailty in people over 60 years of age. Cross-sectional analyses revealed a significant association between nocturia and frailty at follow-up, even though no significant association was identified at baseline. In longitudinal analyses, the prevalence and incidence of frailty at follow-up significantly increased when nocturia was present at baseline. Moreover, we found that frailty, age, and female sex were significantly associated with mortality, while pre-frailty, nocturia, and morbidity burden were not. Overall, our findings contribute to the growing body of evidence suggesting that nocturia may be a relevant risk factor in the development and progression of frailty and reaffirm the known association between frailty and mortality.

### 4.1 Limitations

Before interpreting our results, we identify and discuss the potential sources of bias, imprecision, or confounding that could have affected our findings.

Mortality: the most reliable analysis in this study pertains to the risk factors for death, as it includes all participants in BASE II regardless of their participation in the follow-up. We obtained the needed vital status information from the civil register, thus mitigating the impact of an incomplete follow-up.

Frailty: (1) Selection bias has occurred due to losses to follow-up. Participants not included in the follow-up had been older, more often frail or pre-frail, and tended to have a higher morbidity burden at baseline (**Supplementary Table 1**). Not least, participants deceased after the baseline assessment could not be included in the follow-up. However, death as competing risk factor could not be accounted for in the analysis, since the BASE-II design relied on two cross-sectional surveys without time-to-event data for frailty. Consequently, our analysis may yield biased estimates of the association between nocturia and frailty, as it does not consider the impact of mortality on the development of frailty. (2) Frailty was assessed at baseline and again at follow-up; however, shorter-term, or even repeated transitions in frailty status might occur. For a more precise approach, it would be desirable to measure frailty at several fixed intervals after baseline, giving a clearer picture of the frailty development in the population.

Variable follow-up times: the heterogeneous follow-up times (**Supplementary Figure 1**) pose a challenge in interpreting the relationship between nocturia and frailty. If the relationship is indeed causal, the varying durations of nocturia exposure could affect the observed impact. This variability in exposure time might lead to inconsistencies in how nocturia influences frailty across participants. Nevertheless, the observation period, with a median of 7.1 years (IQR 6.2-8.7), is a valuable strength of this study, allowing potential causal factors sufficient time to manifest their effects at least partially.

Self-reported data: many variables in this study, including nocturia, sleep duration, and sleep quality, were self-reported. These self-reported data introduce the possibility of recall and information bias, potentially leading to misclassification and, consequently, to under- or overestimation of the strength of the association between the exposures and outcome.

Sample size: BASE-II was designed to identify and characterize factors associated with ‘healthy’ vs. ‘unhealthy’ ageing [15] and participants were of the above-average health (reflected in the low morbidity burden) and education [15], [16]. Consequently, our sample has a low frailty prevalence, limiting the ability to model this subgroup comprehensively. Aside from that, we observed reasonable confidence interval widths in most of our analyses.

Generalizability: it has been shown that nocturia is more prevalent in ethnic minorities and people with lower socioeconomic status [30], while BASE-II participants were mostly of European ancestry and educated above average. This selection bias limits the generalizability of our findings to populations other than the source population. The comparatively low prevalence of frailty and nocturia at both time points may also be explained by the above-average health (reflected in the low morbidity burden) and education of BASE-II participants [16].

### 4.2 Interpretation Nocturia in BASE II

As expected, the prevalence of pre-frailty, frailty, and nocturia increased over the follow-up time of more than seven years. Nevertheless, compared to other cohorts, the prevalence and incidence of frailty and nocturia were relatively low, both at baseline and follow-up [5],[31],[18]. Interestingly, at both baseline and follow-up, people with nocturia were about equally divided between men and women. This almost equal sex ratio may imply that sex-specific etiologies underlying nocturia, such as benign prostatic hyperplasia or pelvic floor problems, are unlikely to play a major role. Nocturia at baseline was associated with more sleep disturbances and with lower subjective sleep duration and quality, showing consistency with the nocturia phenotype.

#### Nocturia and frailty

It is known that the prevalence of nocturia increases sharply with age, as does frailty [32], [33]. While nocturia is known to be associated with several adverse outcomes, including night falls, fractures, sleep disturbance, reduced quality of life, depression, and increased mortality [18], the longitudinal association between nocturia and frailty was not previously studied [14]. Our results confirmed for the first time that nocturia is associated with an increased risk of *developing* frailty –the prevalence and incidence of frailty at follow-up increased significantly when nocturia was present at baseline.

Taken together, our analyses suggest that the main temporal pathway goes from nocturia to pre-frailty/frailty, as the risk for worsening frailty status over time was consistently increased in the nocturia subgroups (**Table 1**, Figure 2, **Table 3**). Although the reverse pathway (pre-frailty/frailty followed in time by nocturia) may also occur, this could not be determined in our sample due to the small numbers in the frail group at both baseline and follow-up. Our findings have to be confirmed and substantiated in future studies with higher sample size and more frequent measurement dates to study these transitions more comprehensively.

There has been scarce and heterogeneous evidence of the association between nocturia and frailty from cross-sectional studies. While there are also null results, most studies reported some evidence linking nocturia to frailty. For example, Soma et al. showed in a cross-sectional study of 710 individuals that nocturia (≥ 2 episodes per night) was associated with frailty, which was measured in different ways (Fried Frailty Phenotype (OR 2. 15, 95% CI 1.42-3.24, p < 0.001), modified frailty index (OR 2.00, 95% CI 1.30-3.08, p = 0.002) and frailty discriminant score (OR 2.09, 95% CI 1.45-3.02, p < 0.001)[34]. Likewise, our cross-sectional analyses yielded inconsistent results, as there was no evidence of an association between nocturia and frailty status at baseline, whereas such an association was evident at follow-up.

There is further epidemiological evidence of an association between nocturia and frailty. In a recent review [14], we have shown that shift workers are regularly affected by both nocturia and frailty. In this case, the temporal sequence of the occurrence of nocturia and frailty is clear: first, the circadian rhythm is disturbed, then nocturia symptoms start, and then frailty appears. Indeed, sleep disruption and circadian rhythm may be a central mechanistic link between nocturia and frailty in older adults [14]. Circadian rhythm is central to homeostasis, and its disruption affects several physiological processes and signaling pathways, resulting, for example, in metabolic dysfunction and increased inflammation [35], [36], [37]. Taken together, these factors may ultimately increase the likelihood of frailty.

The analogy to incontinence and frailty is obvious, as both conditions often coexist, share pathophysiological factors, have common underlying conditions, and significantly impact on functional status and quality of life. However, the evidence linking incontinence and frailty is likewise thin. For example, Miles et al. concluded that incontinence is associated with an increased risk of more global functional impairment and an important early marker for signaling the onset of frailty among persons who become incontinent after the age of 65 years. Additionally, Coll-Planas et al. have convincingly conceptualized the disablement process from urinary incontinence to late-life disability [38], [39]. This could apply in a similar way to nocturia and frailty: Pre-frailty conditions are, as we see in BASE-II, more prevalent with increasing age. This has to do with the increase in risk factors such as nocturia. If nocturia and other unfavorable, risk-increasing conditions continue and add up, they promote the transition to frailty. Moreover, frailty can concurrently exacerbate nocturia through a feedback loop.

#### Outlook

The present analysis emphasizes that nocturia is a highly relevant clinical problem that requires increased attention and research –not least because people with nocturia experience high levels of suffering [4]. As our data show, nocturia is associated with a significantly increased occurrence of frailty. Nocturia may be a risk factor but also an early sign or precursor of frailty. Since aging is associated with a weakening in the amplitude of the circadian rhythm and appears to be closely intertwined with nocturia and frailty [14], future studies should consider the aspect of circadian rhythmicity and its disruption. Many lifestyle factors and living conditions in old age have the potential to exacerbate a decline of the circadian rhythm, such as loss of daily structure due to retirement, less time outdoors due to difficulty walking, and less locomotor capacity and activity, among others [40]. Thus, appropriate, rhythm-stabilizing or reinforcing interventions, be it timed exercise, diet, light, or sleep situation, have the potential to both reduce nocturia and slow or stop the transition to frailty and could likewise benefit the aging process beyond nocturia and frailty. It will be worthwhile to see whether consistent treatment of nocturia can reverse existing frailty or pre-frailty in patients in sufficiently sized, randomized, controlled studies.

## 5. Conclusions

This study contributes to the growing evidence suggesting that nocturia may be a significant risk factor in frailty progression. Although the present study already provides valuable insights, future research with more targeted designs and fixed follow-up intervals is necessary to deepen our understanding of the hypothesized associations, especially when aiming to study causality. A better understanding will ultimately contribute to a more targeted approach to increase the quality of life in older adults.

## 6. Acknowledgements

### Funding

This article uses data from the Berlin Aging Study II (BASE-II) and the GendAge study, which were supported by the German Federal Ministry of Education and Research under grant numbers #01UW0808; #16SV5536K, #16SV5537, #16SV5538, #16SV5837, #01GL1716A and #01GL1716B. Yulia Komleva received funding by the Clinician Scientist Program of the Deutsche Forschungsgemeinschaft (DFG).

### Data availability

Due to concerns for participant privacy, data are available only upon reasonable request. Please contact the scientific coordinator Ludmila Müller at for additional information.

## 7. Authors’ contributions

MK had the idea for the work. ID and EST provided the data. CM, JM, and MK carried out the analyses. MK, CM, JM and ID interpreted the results. VV produced the alluvial plot. MK took the lead in writing the manuscript together with CM and JM, with valuable input from MG, YK, ID, VV, and EST.

## Data Availability

All data produced in the present study are available upon reasonable request to the authors.

