## Supplementary material for "Nocturia as a Risk Factor for Developing Frailty in Older Adults: Results of the Berlin Aging Study II"

**Supplementary Table 1.** Characteristics of participants included and excluded from the analysis.

|  | **Included**  1083 (64.8%) | **Excluded***  588 (35.2%) | **p-value** |
| --- | --- | --- | --- |
| Age, mean (SD) | 67.8 (3.5) | 69.2 (3.9) | <0.001 |
| Female sex, n (%) | 563 (52.0) | 299 (50.9) | 0.695 |
| Nocturia, n (%) | 254 (23.6) | 168 (28.7) | 0.027 |
| Frailty at baseline,  n (%) |  |  | 0.043 |
| Robust | 700 (70.2) | 329 (63.9) |  |
| Pre-frail | 288 (28.9) | 181 (35.1) |  |
| Frail | 9 (0.9) | 5 (1.0) |  |
| Morbidity index at baseline,  median (IQR) | 0.97 (1.20) | 1.09 (1.28) | 0.069 |
| Fallen in the past 12 months, n (%) | 325 (30.1) | 183 (31.4) | 0.615 |
| Hospitalizations in the past 12 months, n (%) | 183 (16.9) | 107 (18.4) | 0.502 |
| Subjective sleep duration in hours, median (IQR) | 7 (6-8) | 7 (6-8) | 0.782 |
| Poor sleep quality (subj.), n (%) | 171 (17.2) | 85 (15.7) | 0.497 |

*Notes and abbreviations:* Data are presented as mean (standard deviation, SD), number of observations (percentage), or median (IQR = 25th,75th percentile); *excluded due to death or loss to follow-up.

**Supplementary Table 2.** Participants’ characteristics at baseline and follow-up, stratified by nocturia status (n=1083).

| **Baseline*** | | | | | | | | | |
| --- | --- | --- | --- | --- | --- | --- | --- | --- | --- |
|  | | | **No Nocturia**  821 (75.8%) | **Nocturia***  254 (23.5%) | **Missings**  8 (0.7%) | | | **p-value** | |
| Age, years | | | 67.6 (3.5) | 68.5 (3.3) | 0 | | | <0.001 | |
| Female sex, % | | | 438 (53.3) | 122 (48.0) | 0 | | | 0.158 | |
| Frailty at T0, % | | |  |  | 86 | | | 0.633 | |
| - Robust | | | 539 (70.9) | 158 (68.4) |  | | |  | |
| - Pre-frail | | | 215 (28.3) | 70 (30.3) |  | | |  | |
| - Frail | | | 6 (0.8) | 3 (1.3) |  | | |  | |
| Fallen in the past 12 months, % | | | 250 (30.6) | 74 (29.1) | 4 | | | 0.723 | |
| Hospitalizations in the past 12 months, % | | | 122 (14.9) | 61 (24.0) | 1 | | | 0.001 | |
| Morbidity index | | | 0.91 (1.16) | 1.15 (1.32) | 97 | | | <0.001 | |
| Subjective sleep duration, hours | | | 7 (6.5-8) | 7 (6-8) | 91 | | | 0.015 | |
| Poor sleep quality (subj.), % | | | 104 (13.8) | 67 (28.8) | 91 | | | <0.001 | |
| **Follow-up**** | | | | | | | | | |
|  | **No Nocturia**  656 (60.6%) | | | **Nocturia***  423 (39.1%) | | **Missings**  4 (0.4%) | **p-value** | | |
| Age, years | | 75.3 (3.7) | | 76.1 (3.8) | | 0 | <0.001 | | |
| Female sex, % | | | 362 (55.2) | 200 (47.3) | 0 | | | | 0.013 |
| Frailty T1, % | | |  |  | 13 | | | | 0.005 |
| - Robust | | | 330 (50.6) | 170 (41.0) |  | | | |  |
| - Pre-frail | | | 298 (45.7) | 221 (53.3) |  | | | |  |
| - Frail | | | 24 (3.7) | 24 (5.8) |  | | | |  |
| Fallen in the past 12 months, % | | | 131 (25.1) | 106 (30.7) | 215 | | | | 0.084 |
| Hospitalizations in the past 12 months, % | | | 127 (19.8) | 117 (28.0) | 19 | | | | 0.002 |
| Morbidity index | | 1.32 (1.53) | | 1.52 (1.55) | | 143 | 0.058 | | |

*Notes and abbreviations:* *Nocturia at baseline defined as ≥2 episodes/night; ** nocturia at follow-up; Data are presented as mean (standard deviation), number of observations (percentage), or median (IQR = 25^th^,75^th^ percentile).

**Supplementary Table 3**. Participants’ characteristics at baseline and follow-up, stratified by frailty status (n=1083).

| **Baseline*** | | | | | |
| --- | --- | --- | --- | --- | --- |
|  | **Robust**  700 (64.6%) | **Pre-frail**  288 (26.6%) | **Frail**  9 (0.8%) | **Missings**  86 (7.9%) | **p-value** |
| Age, years | 67.60 (3.29) | 68.21 (3.92) | 67.00 (3.97) | 0 | 0.146 |
| Female sex, % | 355 (50.7) | 155 (53.8) | 5 (55.6) | 0 | 0.656 |
| Nocturia, % | 158 (22.7) | 70 (24.6) | 3 (33.3) | 8 | 0.663 |
| Fallen in the past 12 months, % | 211 (30.2) | 90 (31.2) | 2 (22.2) | 4 | 0.823 |
| Hospitalization in the past 12 months, % | 120 (17.1) | 45 (15.6) | 2 (22.2) | 1 | 0.766 |
| Morbidity index | 0.88 (1.10) | 1.16 (1.33) | 1.67 (1.41) | 97 | 0.005 |
| Subjective sleep duration, hours | 7 (6.5-8) | 7 (6-8) | 8.3 (7.3-8.9) | 91 | 0.003 |
| Poor sleep quality (subj.), % | 93 (14.3) | 64 (24.3) | 1 (16.7) | 91 | 0.001 |
| **Follow-up**** | | | | | |
|  | **Robust**  502 (46.4%) | **Pre-frail**  520 (48.0%) | **Frail**  48 (4.4%) | **Missings**  13 (1.2%) | **p-value** |
| Age, years | 75.09 (3.76) | 75.96 (3.60) | 77.45 (4.55) | 0 | <0.001 |
| Female sex, % | 254 (50.6) | 276 (53.1) | 28 (58.3) | 0 | 0.497 |
| Nocturia, % | 170 (34.0) | 221 (42.6) | 24 (50.0) | 4 | 0.005 |
| Fallen in the past 12 months, % | 95 (24.4) | 127 (29.4) | 12 (32.4) | 215 | 0.207 |
| Hospitalization in the past 12 months, % | 104 (21.1) | 124 (24.3) | 15 (31.2) | 19 | 0.187 |
| Morbidity index | 1.22 (1.41) | 1.45 (1.54) | 2.62 (2.16) | 143 | <0.001 |

*Notes and abbreviations:* *Frailty at baseline; ** Frailty at follow-up; Data are presented as mean (standard deviation), numbers(proportions), or median(IQR = 25^th^,75^th^ percentile).

**Supplementary figure 1**.

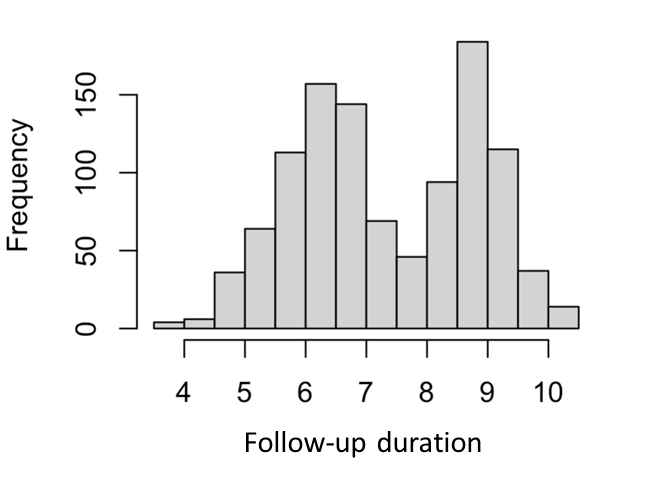

**Supplementary figure 1:** Bi-modal distribution of observed follow-up duration (in years).

**Supplementary figure 2a.**

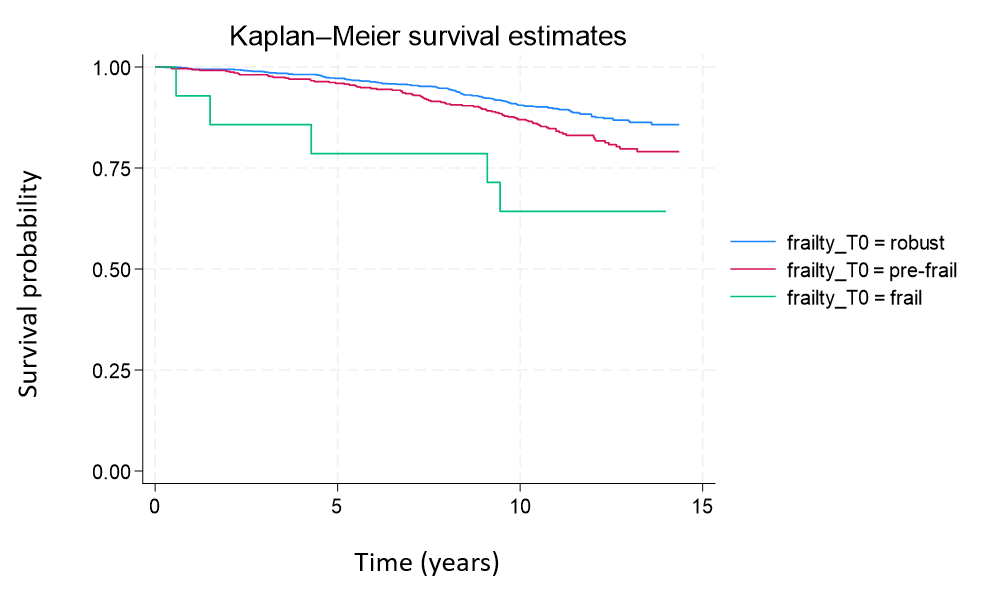

**Supplementary figure 2b.**

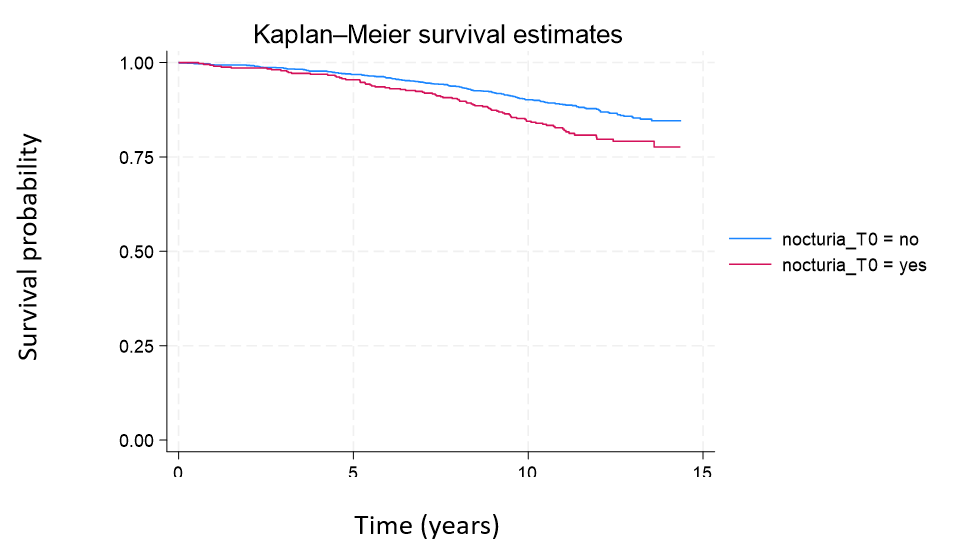

**Supplementary figures 2a-b:** Kaplan-Meier survival estimates for BASE-II (n=1671) by a) frailty status and b) nocturia.
